# Comparing Clinicopathologic Features and Outcomes Between Young Children, Adolescents, and Adults with Rhabdomyosarcoma at a Single Institution

**DOI:** 10.1101/2025.01.13.25320489

**Authors:** Karla Boyd, Nalin Leelatian, Raffaella Morotti, Hari Deshpande, Emily Christison-Lagay, Sarah McCollum, Veronika Shabanova, Farzana Pashankar, Juan C. Vasquez

## Abstract

**Background:** There are limited data comparing clinical and pathologic features and outcomes of rhabdomyosarcoma (RMS) between young children, adolescent/young adults (AYAs), and adults.

**Design/Methods:** We performed a retrospective chart review of patients with RMS treated at our institution between 2000 and 2017. We compared clinical and pathologic features, treatment modalities, and outcomes among three different age groups (young children: <15 years of age, AYA: 15-39 years of age, and older adults: >39 years of age).

**Results:** Among 65 patients, 33 (50.8%) were young children, 14 (21.5%) were AYAs, and 18 (27.7%) were older adults. Head and neck was the most common tumor site among young children, AYAs tumor sites were variable, while extremities were the most frequent location for adults. AYAs were more likely to have stage 4 disease at presentation (64.3% vs. 21.2% in children, p=0.01, and vs. 27.8% in adults, p=0.08). Similarly, clinical group and risk classifications were higher among AYAs as compared to the other two age groups. Tumors in children were alveolar and embryonal histology, while AYAs primarily developed alveolar RMS. Older adults had tumors with diverse histologies, including pleiomorphic and spindle cell. Children and AYAs received multimodal therapy, but a subset of adults did not. Five-year overall survival (OS) varied by age at diagnosis (p=0.001), with highest OS among young children (81.8%) following by adults (50.0%), with the lowest OS among AYAs (14.29%).

**Conclusion:** This study highlights the need for ongoing collaboration between pediatric and adult multidisciplinary teams to improve outcomes for patients with RMS across the age spectrum.

## Introduction

Rhabdomyosarcoma (RMS), a neoplasm of skeletal muscle, is the most common soft tissue sarcoma in children. It accounts for less than 5% of adult sarcomas, and half of all cases are found in children less than 6 years of age. While 5 year overall survival (OS) rates in children have improved over the past 30 years to more than 60%, 5-year OS in adolescents and adults remains closer to 25-30%, with the exception of young adolescent patients with low-stage disease.^1^ This difference appears to be caused in part by higher risk disease in these age groups, with an increased frequency of alveolar histology and *FOXO 1* fusion positivity, less favorable primary tumor locations, more advanced disease at presentation, as well as treatment modality.^2^ However, most studies looking at the effect of age on RMS compare either children with adults or children with AYAs, but do not look at all three groups together. There is also variability in the definition of these groups, with adults defined in some studies as patients >18 years of age but in others as > 40 years.

In addition to presenting with different tumor biology compared to young children, adolescent/young adults (AYAs) are underrepresented on prospective clinical trials^2^. Treatment of RMS in AYAs is variable, determined specialty by either an adult or pediatric oncologist and dependent on institutional practices. Adult patients are less likely to be treated with multimodal therapy that has been shown to result in improved survival outcomes and have lower enrollment on clinical trials.^3^ Because older patients have worse survival outcomes than young children, current Children’s Oncology Group (COG) guidelines use an age cutoff of 10 years to determine if patients with fusion-negative, stage 4 disease should be treated as intermediate or high risk.^5^

The aim of this study was to examine the differences in presentation, treatment, and outcomes between those diagnosed with RMS as children, AYAs, and adults. We hope that a better understanding of the biological features and outcomes of RMS across different ages will highlight the need for improved cooperation between pediatric and adult multidisciplinary teams in delivering coordinated care and designing future clinical trials to improve outcomes.

## Methods

### Patient Population

This study was conducted at Smilow Cancer Center/Yale New Haven Hospital, a large teaching hospital that provides care to patients with cancer across the lifespan. The hospital maintains a robust pathology database of all patients who have had a pathology specimen from diagnostic biopsy or resection. From this database, we identified patients who were diagnosed with rhabdomyosarcoma between 2000 and 2017. After removing duplicate patient entries, the electronic medical record (EMR) was reviewed, and clinical data was extracted. This included age at diagnosis, sex, primary tumor location, histological subtype of RMS, *PAX/FOXO1* fusion status, TNM stage, clinical group, treatment modalities, dates of any relapse, secondary cancers, death, and last follow-up. Favorable tumor locations were defined as non-parameningeal head and neck (including orbit), non-prostate-non-bladder genitourinary (GU), and biliary. All other locations were classified as unfavorable. This information was then used to derive COG risk group using current risk stratification guidelines^5^. COG risk groups were applied to all patients, regardless of age.

The COG standard of care for RMS uses multimodal therapy comprised of chemotherapy with a vincristine/dactinomycin/cyclophosphamide backbone, radiotherapy in the majority cases, and surgical resection when feasible. There is no definitive national guideline for the treatment of RMS in adults, possibly due to its rarity in this population, and pediatric regimens are often used^21^. In this analysis, we extracted treatment modalities from the EMR to determine if the approach was in line with COG standard of care. This study was submitted to and approved by the Yale Institutional Review Board (IRB).

### Statistical analysis

Patients were categorized by age at RMS diagnosis as children, adolescent/young adults (AYAs), or adults (ages <15, 15-39, and >39, respectively) for the purpose of statistical comparisons. Age at diagnosis, sex, and clinical characteristics were described using summary statistics such as counts (percentage) and median (25^th^, 75^th^ percentiles). Differences in these characteristics between age groups were tested using Chi-square, Fisher’s exact, Wilcoxon Rank Sum test, as appropriate. Overall, 5-year survival (OS) curves and event-free 5-year survival (EFS) curves were estimated using Kaplan-Meier survival analysis and compared by age group using the log-rank test. Since all pair-wise comparisons among the age groups at diagnosis were *a priori* independent hypotheses of interest, we did not adjust for multiple comparisons^7^, Regression analysis based on the Cox proportional hazards model was used to examine which predictors were associated with the differences in 5-year overall (OS) and event-free (EFS) survival time in the full sample and stratified by age. Proportional hazards assumption was evaluated using the standardized score process, which is based on the cumulative sums of martingale-based residuals^13^. Results were summarized as estimated proportion or percent 5-year OS and EFS, and unadjusted hazard ratios (HR) for survival (not event or mortality) with surrounding 95% Confidence Intervals (95%CI). All analyses were completed in SAS version 9.4 (Cary, NC).

## Results

In total 65 patients were identified, including 33 young children (median age 5 years, range 1.1-13), 14 AYAs (median age 19 years, range 15-37) and 18 older adults (median age 59 years, range 45-95). Follow-up within 5 years (time to death or last follow-up date) ranged from 1 to 60 months (median 34 months, IQR 12-60 months), and was the longest for children, followed by adults and then AYAs (Table 1).

**Table 1:** Patient Characteristics.

|  | <u>Children <15yrs,</u><br><u>n=33</u> |  | <u>AYA 15-39yrs,</u><br><u>n=14</u> |  | <u>Adults 40+ yrs,</u><br><u>n=18</u> |  | **Total, n=65** |  | **P value** |
| --- | --- | --- | --- | --- | --- | --- | --- | --- | --- |
| **Follow up within 5 years in months, median (25<sup>th</sup>, 75<sup>th</sup> percentile)** | 60.0 (21.0, 60.0) |  | 16.0 (8.0, 25.0) |  | 26.5 (9.0, 60.0) |  | 34.0 (12.0, 60.0) |  | 0.004 |
|  | Number | (%) | Number | (%) | Number | (%) | Number | (%) |  |
| **Sex** |  |  |  |  |  |  |  |  | 0.49 |
| Female | 13 | 39.4 | 8 | 57.1 | 7 | 38.9 | 28 | 43.1 |  |
| Male | 20 | 60.6 | 6 | 42.9 | 11 | 61.1 | 37 | 56.9 |  |
| **Site** |  |  |  |  |  |  |  |  | 0.10 |
| Favorable<sup>1</sup> | 13 | 39.4 | 1 | 7.1 | 5 | 27.8 | 19 | 29.3 |  |
| Unfavorable | 20 | 60.6 | 12 | 85.7 | 13 | 72.2 | 45 | 69.2 |  |
| Unknown | 0 | 0 | 1 | 7.1 | 0 | 0 | 1 | 1.5 |  |
| **Pathological Subtype** |  |  |  |  |  |  |  |  | <0.001 |
| Alveolar | 16 | 48.5 | 10 | 71.4 | 2 | 11.1 | 28 | 43.1 |  |
| Embryonal | 11 | 33.3 | 1 | 7.1 | 1 | 5.6 | 13 | 20.0 |  |
| Spindle Cell/Sclerosing | 1 | 3.0 | 1 | 0 | 3 | 16.7 | 5 | 7.7 |  |
| Botryoid | 4 | 12.1 | 0 | 0 | 0 | 0 | 4 | 6.2 |  |
| Pleomorphic | 0 | 0 | 0 | 0 | 9 | 50.0 | 9 | 13.9 |  |
| Mixed | 1 | 3.0 | 0 | 0 | 2 | 11.1 | 3 | 4.6 |  |
| Poorly Differentiated | 0 | 0 | 1 | 7.1 | 0 | 0 | 1 | 1.5 |  |
| Unknown | 0 | 0 | 1 | 7.1 | 1 | 5.6 | 2 | 3.1 |  |
| **FOXO1/PAX Fusion Status** |  |  |  |  |  |  |  |  | 0.01 |
| Positive | 2 | 6.1 | 7 | 50.0 | 1 | 5.6 | 5 | 7.7 |  |
| Negative | 5 | 15.2 | 0 | 0 | 0 | 0 | 10 | 15.4 |  |
| Unknown | 26 | 78.8 | 7 | 50.0 | 17 | 94.4 | 50 | 76.9 |  |
| **Stage** |  |  |  |  |  |  |  |  | 0.08 |
| I | 10 | 30.3 | 1 | 7.1 | 2 | 11.1 | 13 | 20.0 |  |
| II | 4 | 12.1 | 0 | 0 | 3 | 16.7 | 7 | 10.8 |  |
| III | 11 | 33.3 | 4 | 28.6 | 8 | 44.4 | 23 | 35.4 |  |
| IV | 7 | 21.2 | 9 | 64.3 | 5 | 27.8 | 21 | 32.1 |  |
| Unknown | 1 | 3.0 | 1 | 0 | 0 | 0 | 1 | 1.54 |  |
| **Clinical Group** |  |  |  |  |  |  |  |  | 0.03 |
| **1** | 6 | 18.2 | 1 | 7.1 | 4 | 22.2 | 11 | 16.9 |  |
| **2** | 2 | 6.1 | 1 | 7.1 | 2 | 11.1 | 5 | 7.7 |  |
| **3** | 14 | 42.4 | 1 | 7.1 | 4 | 22.2 | 19 | 29.2 |  |
| **4** | 10 | 30.3 | 11 | 78.6 | 3 | 16.7 | 24 | 36.9 |  |
| **Unknown** | 1 | 3.0 | 0 | 0 | 5 | 27.8 | 6 | 9.2 |  |
| **COG Risk Stratification** |  |  |  |  |  |  |  |  | 0.14 |
| **Low risk** | 10 | 30.3 | 1 | 7.1 | 5 | 27.8 | 16 | 24.6 |  |
| **Intermediate Risk** | 12 | 36.4 | 2 | 14.3 | 4 | 22.2 | 18 | 27.7 |  |
| **High Risk** | 10 | 30.3 | 9 | 64.3 | 5 | 27.8 | 24 | 36.9 |  |
| **Unknown** | 1 | 3.0 | 2 | 14.3 | 4 | 22.2 | 7 | 10.8 |  |
| **COG Risk High vs Other** |  |  |  |  |  |  |  |  | 0.02 |
| **Low or Intermediate Risk** | 22 | 66.7 | 4 | 28.6 | 13 | 72.2 | 39 | 60.0 |  |
| **High Risk** | 10 | 30.3 | 10 | 71.4 | 5 | 27.8 | 29 | 38.5 |  |
| **Unknown** | 1 | 3.0 | 0 | 0 | 0 | 0 | 1 | 1.5 |  |
| **Treatment** |  |  |  |  |  |  |  |  | <0.001 |
| **Surgery alone** | 0 | 0 | 0 | 0 | 6 | 33.3 | 6 | 9.23 |  |
| **Chemotherapy Alone** | 1 | 3.0 | 3 | 21.4 | 1 | 5.6 | 5 | 7.7 |  |
| **Surgery/Chemotherapy** | 5 | 15.2 | 1 | 7.1 | 2 | 11.1 | 8 | 12.3 |  |
| **Radiation/Chemotherapy** | 14 | 42.4 | 9 | 64.3 | 3 | 16.7 | 26 | 40.0 |  |
| **Surgery/Radiation/Chemo** | 10 | 30.3 | 1 | 7.1 | 6 | 33.3 | 17 | 26.1 |  |
| **Unknown** | 3 | 9.1 | 0 | 0 | 0 | 0 | 3 | 4.6 |  |

Abbreviations: COG= Children's Oncology Group
1: Non-parameningeal head and neck (including orbit), non-prostate-non-bladder genitourinary (GU), and biliary

Histology varied by age group (Table 1). Tumors in young children exhibited either alveolar (ARMS) or embryonal (ERMS) histology, and four of the ERMS tumors were of the botryoid subtype. The majority of the AYAs’ tumors were of alveolar histology, with only one embryonal, one spindle cell/sclerosing, and one poorly differentiated. In adults, there was a smaller proportion of ERMS and ARMS tumors; half of all tumors were classified as pleiomorphic (PRMS), a subtype which was not seen in either of the other age groups. Additioinaly among those diagnosed with RMS as adults, there were also three spindle cell/sclerosing (SCSR) and two mixed histology RMS.

Primary tumor location was heterogenous in all age categories (Fig. 1a). However, head and neck tumors were seen most frequently in young children (36.3%), and older adults were the most likely to have extremity tumors (50.0 %) (p=0.01). AYAs did not show a propensity for any specific location, but they did have the highest rate of tumors in unfavorable sites (85.7%), whereas older adults had an unfavorable site rate of 72.2%, and young children had the lowest rate (60.6%) (p=0.10, Table 1).

**Figure 1.**
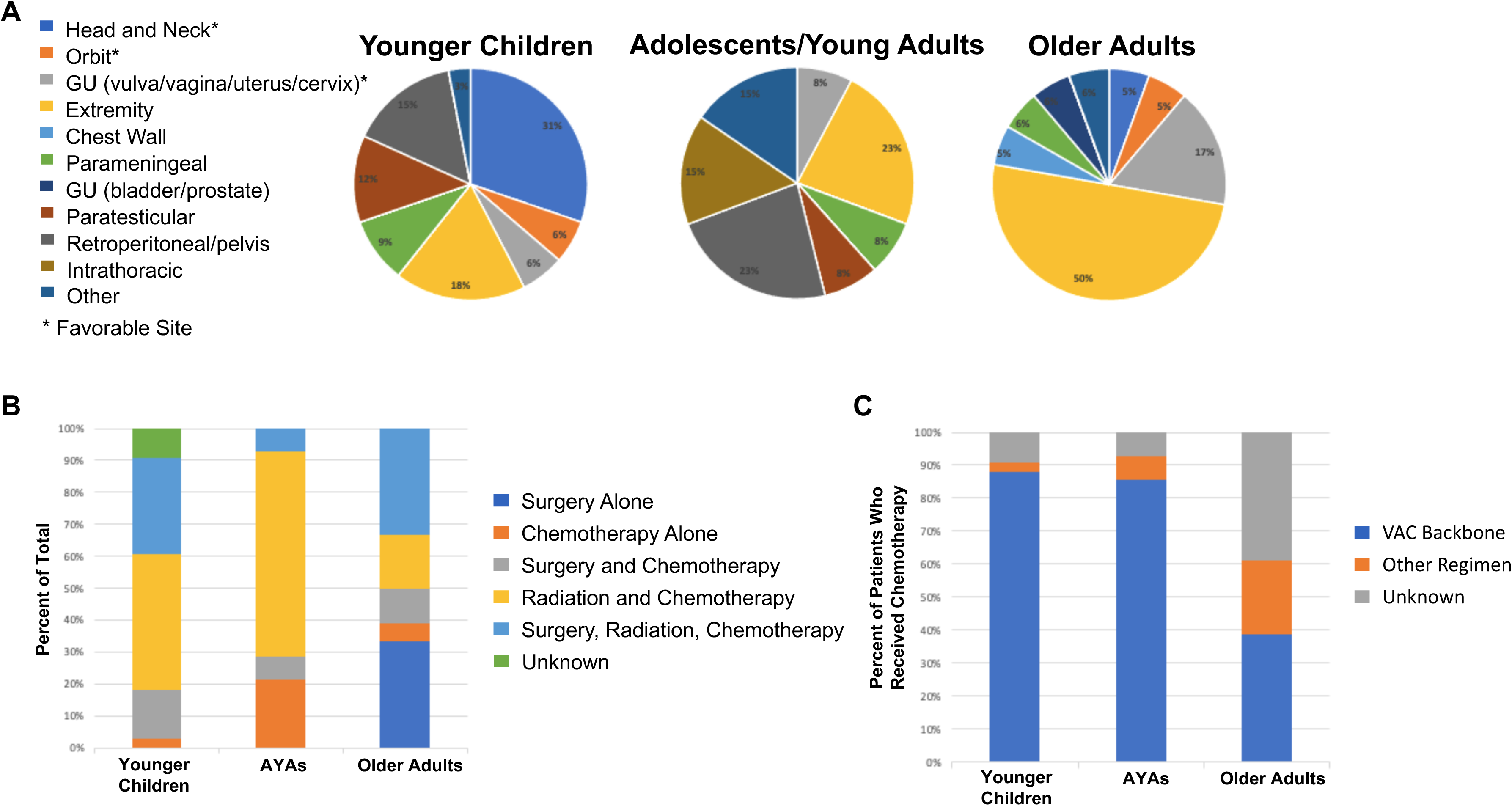
(A) Tumor primary locations among children, AYA, and adults. (B) Treatment modalities received by patients within each age group. (C) Chemotherapy treatment with VAC backbone vs another regimen. Percentages are out of total patients within that age group. P-values were obtained using the Fisher’s exact test.

At presentation, AYA patients had the highest prevalence of advanced stage disease, with 64.3% having metastatic disease at diagnosis as compared to only 21.2% of children and 27.8% of adults (p=0.08) (Table 1). Applying the COG risk stratification^6^ to this cohort of patients, we found that AYAs overall presented with higher risk disease (64.3% high-risk) (Table 1). Young children and older adults had much lower rates of high-risk disease, with only 27-30% falling into high-risk categories (p=0.02).

We examined *PAX/FOXO1* fusion status in our cohort of patients (Table 1) and found that the majority of young children (78.8%) did not have *FOXO1* fusion testing performed, likely reflecting the preponderance of embryonal histology. Moreover, of young children with alveolar histology, only two out of seven tested, or 28.6%, were positive for *PAX/FOXO1* fusion. Among AYAs, half of tumors were tested for fusion status, and all were positive for PAX/FOXO1. With the exception of one tumor testing positive, tumors at presentation in older adults were not tested for PAX/FOXO1 (Table 1).

AYAs and children were more likely to receive multimodal treatment (chemotherapy plus surgery and/or radiation) (78.6% and 87.9%, respectively) (Fig. 1b). Among older adults, 33.3% had surgery alone, but the majority (61.1%, p<0.001) did have multimodal treatment. Only 38.9% of adult patients were treated with regimen consisting of a VAC backbone, which is the basis of pediatric protocols, compared to 71.8% of AYAs and 85.7% of young children (p=0.02, Fig. 1c). Other regimens used included doxorubicin/vincristine, doxorubicin/ifosphamide, doxorubicin/ifosphamide/dacarbazine, and gemcitabine and docetaxel.

Lastly, we assessed survival outcomes by age group. Young children had the highest five-year survival (OS=81.8% and EFS=75.8%) (Fig. 2). AYAs’ five-year OS and EFS was lowest of all three groups, at 14.3% (OS: p=0.0001 vs. young children and p=0.04 vs. adults, and EFS: p<0.0001 vs. young children and p=0.04 vs. adults); although adults’ survival numbers were also lower than young children’s (OS 50.0%, p=0.01 vs. children, and EFS 50.0% p=0.04 vs. children). In the unadjusted analyses on the total sample, having higher clinical group, higher cancer stage, COG high risk stratification, presence of metastasis and single treatment modality were negatively associated with 5-years OS and EFS, while receiving VAC therapy and tumor in a favorable site were positively associated with OS and EFS (Table 2). Young children were more likely to have a more favorable tumor location (39.4%, vs 7.1% among AYAs and 27.8% among older adults), and this study lacked to the power to separate tumor site or young age by as the primary factor influencing OS by multivariable regression. We suspect that they both contribute.

**Figure 2:**
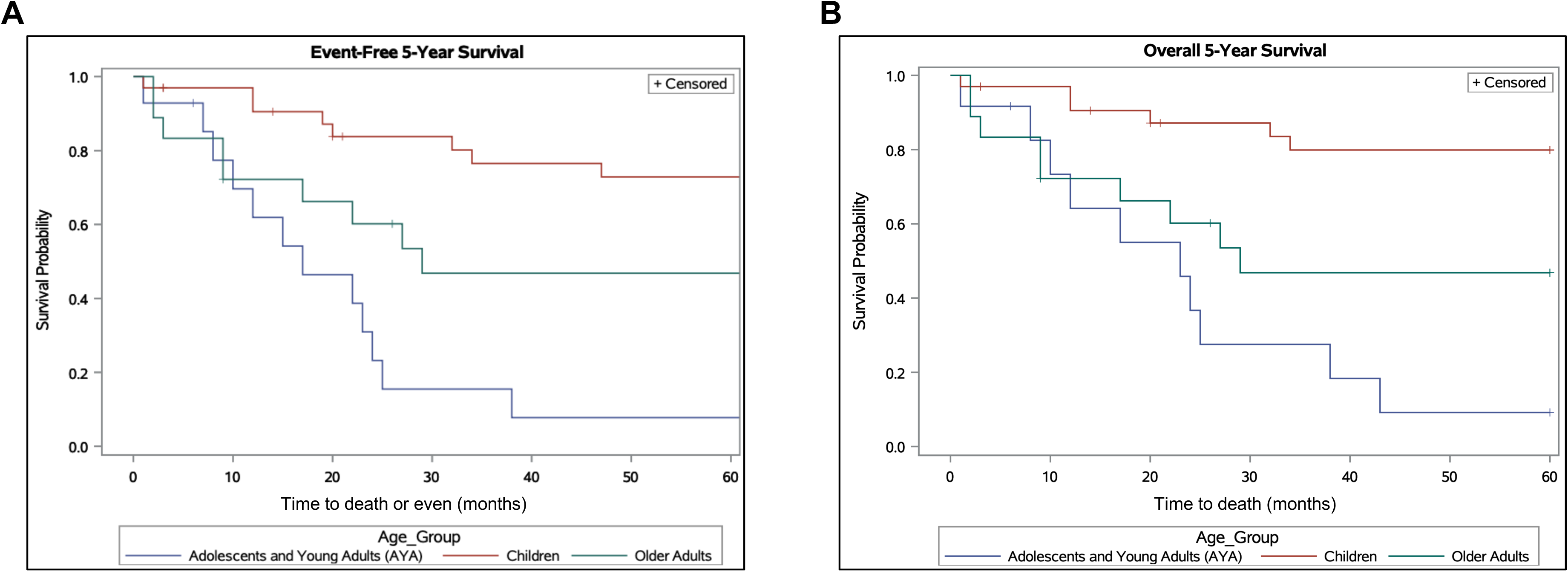
Five-year OS (A) and EFS (B) of patients separated by age group.

**Table 2.**
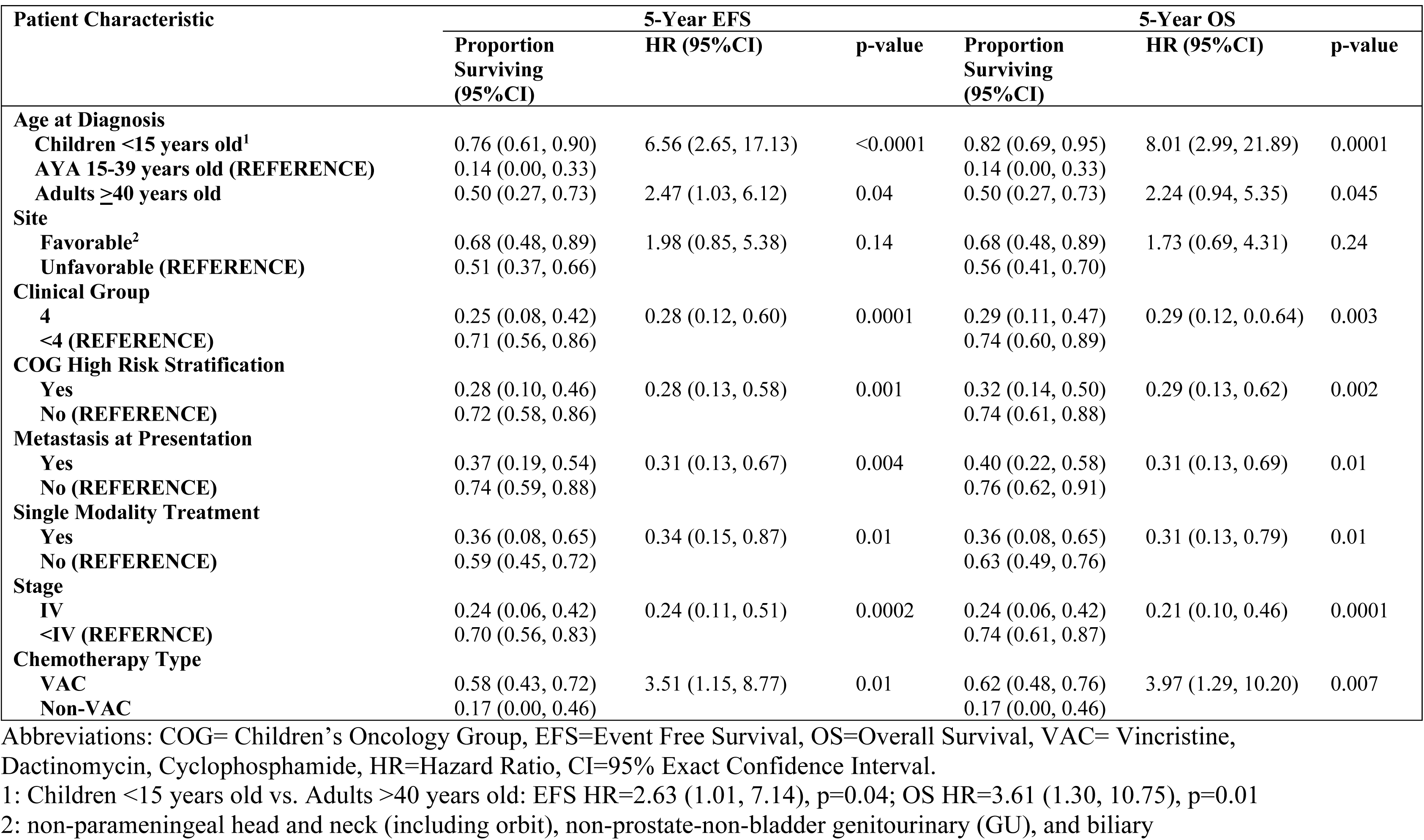
Unadjusted Associations Between Survival and Patient Characteristics.

The difference in 5-year OS and EFS persisted between children and AYAs in all adjusted analyses; with differences in 5-year OS and EFS between children and older adults associated with a higher prevalence of single treatment modality among adults (adjusted for single treatment modality HR=2.8 [p=0.10] for children vs. adults in OS, adjusted HR=2.8 [p=0.25] in EFS) and by the lower prevalence of treatment regimen with VAC (adjusted for VAC regimen HR=1.9 [p=0.38] for children vs. adults in OS, adjusted HR=1.3 [p=0.70] in EFS); the differences in both EFS and OSS between AYAs and older adults were attributable to the more advanced presentation at diagnosis among AYAs (p>0.05 for HR of AYA vs. older adults when adjusted for either clinical group, COG high risk stratification, cancer stage at diagnosis or metastatic at presentation). In the stratified by age group at diagnosis regression analyses, the strongest negative predictor for 5-year survival among AYAs was single treatment modality (HR=0.17 [95%CI 0.03-0.93, p=0.03] for both 5-year OS and EFS); and among both children and adults the strongest negative predictors of 5-year survival were high COG risk (children: HR=0.23 [95%CI 0.03-1.0, p=0.05] for OS, and HR =0.24 [95%CI 0.05-0.98, p=0.045] for EFS; adults: HR=0.24 [95%CI 0.06-1.0, p=0.05] for both OS and EFS) and stage IV cancer at diagnosis (children: HR=0.24 [95%CI 0.04-1.1, p=0.06] for OS; adults: HR=0.24 [95%CI 0.06-0.99, p=0.045] for both OS and EFS).

## Discussion

Although it is most commonly seen among children less than six years of age, rhabdomyosarcoma (RMS) can develop at any age point in a patient’s life. Adults as a group have worse outcomes than children, with little to no improvement in survival outcomes over the past 50 years.^8,9^ Most of the studies examining RMS outcomes and age have compared either children to adults or children to adolescent/young adults (AYAs), and a variety of age cutoffs have been applied. There is much less data examining differences between all three of these age groups. One study by Fischer et al from 2018 looking at National Cancer Database data from 1998-2012 found that five-year OS was best among children (74%) and worst for adults (35%), with AYAs in between (50%). They also showed that multimodal therapy was independently associated with better OS in the adult and AYA groups.^10^ Our single-institution study was smaller but looked at more recent patients (2000-2016) and we had access to granular data such as specific chemotherapy regimens and tumor locations through our medical record system.

Within our cohort, AYAs had poorer outcomes than either older or younger patients, with lower OS and EFS. This was despite being more likely to receive multimodal treatment and having the highest rate of pediatric protocol utilization. A recent position paper from the European Society for Medical Oncology (ESMO) and the European Society for Paediatric Oncology (SIOPE) noted that AYAs with cancer have not had the survival gains that we have seen among children and adults over the past few decades, and emphasized a need to correct this.^11,12,14^ A 2016 review of the Surveillance, Epidemiology, and End Results Database (SEER) data came to a similar conclusion.^14^ There have been several proposed reasons for this, including more aggressive tumor biology, less aggressive treatment, AYAs not being referred to specialized care centers, delays in diagnosis, and lack of inclusion on clinical trials.^15^ Others have noted that AYAs and adults have better survival rates when treated on pediatric RMS protocols and with multimodal therapy^3,10,15^. In this study, many of those concerns do not apply. All patients were at the same specialized center. The AYAs had the highest rate of clinical trial participation and were the most likely to be treated using pediatric protocols, and 78.57% of them received multimodal treatment, which is similar to children (87.88%) and greater than adults (61.11%).

Tumor biology may also have contributed to poorer outcomes for AYAs. They had a higher proportion of alveolar histology (71.43%) than children (48.48%). We have learned over the past two decades that *PAX/FOXO1* fusion status is a better indicator of tumor aggressiveness than histology, and testing is now standard of care and incorporated into the newest COG risk stratification.^17^ These patients were diagnosed between 2000 and 2017, and only those samples obtained from 2009 onward were consistently tested for PAX/FOXO 1 fusions. This included seven children, two of whom were fusion-positive and five of whom were fusion negative. The one adult who was tested was positive, and seven of the eight AYAs were positive. The aggressive nature of so many of the AYAs tumors likely contributed to their lower OS and EFS.

Treatment modality, in particular use of single vs. multimodal therapy, was found on regression analysis to be associated with differences in OS and EFS. Some of the AYAs only received chemotherapy, with no local control. These patients all had widely metastatic disease at diagnosis, including a primary tumor or metastases to the abdomen/pelvis. Several adults were treated with surgery alone. One of them had localized PRMS, for which surgery is the standard of care. However, two had metastatic PRMS, two had a mixed-histology tumor (one metastatic, the other localized), and another tumor had unknown histology.

As noted earlier, there is literature suggesting that more access to clinical trials is needed to improve AYA outcomes, and four of our AYA patients did in fact participate in a trial. The trial was ARST08P1, which tested the use of cixutumumab or temozolomide in addition to standard RMS chemotherapy regimens. Unfortunately, this trial did not show improvement in outcomes with either of these agents.^18^

There has been emerging literature pointing to decreased survival rates experienced by people of color with cancer. A recent study looking at patients with AML found that structural racism mediates much of these survival disparities.^19^ Another paper by Munnikhuysen et al looked at RMS patients specifically and found that although Non-Hispanic Black and Hispanic patients tended to present with more high-risk disease, there was no difference in overall or event-free survival by ethnic group.^20^ Our cohort only included 23 people of color, which was not enough to draw strong conclusions.

This study had several limitations. The total number of patients is small, potentially limiting inferences from our regression analyses, but we presented confidence intervals around our estimates to stress the coverage of mostly protective or detrimental effects of a predictor, and we did not solely rely on p-values. This study only looks at patients who live in or near Connecticut, and as noted above, there is a lack of racial diversity in this cohort. Only a subset of patients underwent *PAX/FOXO1* fusion, making it difficult to determine differences in the molecular profile of tumors by age and its effect on outcome.

AYAs are a unique group with biological, social and emotional needs that are different than either younger or older patients. Better understanding of the reasons behind their poorer oncologic outcomes is necessary to improve their care. Moreover, our findings underscore the vital need for ongoing cooperation between pediatric and adult multidisciplinary teams in the care of RMS and the implementation of future clinical trials to improve outcomes in RMS across the age spectrum.

## Data Availability

All data produced in the present study are available upon reasonable request to the authors

## Abbreviations

RMS: Rhabdomyosarcoma
AYA: dolescent/Young Adult
ERMS: Embryonal rhabdomyosarcoma
ARMS: Alveolar rhabdomyosarcoma
PRMS: Pleiomorphic Rhabdomyosarcoma
SCSR: Spindle Cell/Sclerosing Rhabdomyosarcoma
OS: Overall survival
EFS: Event-free survival
EMR: Electronic Medical Record
GU: Genitourinary
ESMO: European Society for Medical Oncology
SIOPE: European Society for Paediatric Oncology
TNM: Tumor/Node/Metastasis (Internationally Used Cancer Staging System)

## Acknowledgements

JCV is funded in part by the Robert Wood Johnson Harold Amos Medical Faculty Development Program and the Fund to Retain Clinical Scientists at Yale, sponsored by the Doris Duke Charitable Foundation award #2015216, and the Yale Center for Clinical Investigation.

## References

1. Leiner J, Le Loarer F. The current landscape of rhabdomyosarcomas: an update. Virchows Arch. 2020;476:97–108

2. Skapek SX, Ferrari A, Gupta AA, Lupo PJ, Butler E, Shipley J, Barr FG, Hawkins DS. Rhabdomyosarcoma. Nat Rev Dis Primers. 2019;5:1

3. Gerber NK, Wexler LH, Singer S, Alektiar KM, Keohan ML, Shi W, Zhang Z, Wolden S. Adult rhabdomyosarcoma survival improved with treatment on multimodality protocols. Int J Radiat Oncol Biol Phys. 2013;8658–63

4. Alken S, Owens C, Gilham C, Grant C, Pears J, Deady S, O’Marcaigh A, Capra M, O’Mahony D, Smith O, Walsh PM. Survival of childhood and adolescent/young adult (AYA) cancer patients in Ireland during 1994-2013: comparisons by age. Ir J Med Sci. 2020;189:1223–1236

5. Haduong JH, Heske CM, Allen-Rhoades W, Xue W, Teot LA, Rodeberg DA, Donaldson SS, Weiss A, Hawkins DS, Venkatramani R. An update on rhabdomyosarcoma risk stratification and the rationale for current and future Children’s Oncology Group clinical trials. Pediatr Blood Cancer. 2022;69:e29511

6. Crane JN, Xue W, Qumseya A, Gao Z, Arndt CAS, Donaldson SS, Harrison DJ, Hawkins DS, Linardic CM, Mascarenhas L, Meyer WH, Rodeberg DA, Rudzinski ER, Shulkin BL, Walterhouse DO, Venkatramani R, Weiss AR. Clinical group and modified TNM stage for rhabdomyosarcoma: A review from the Children’s Oncology Group. Pediatr Blood Cancer. 2022;69:e29644

7. Rubin, M. (2021). When to adjust alpha during multiple testing: A consideration of disjunction, conjunction, and individual testing. Synthese, 199, 10969–11000.

8. Egas-Bejar D, Huh WW. Rhabdomyosarcoma in adolescent and young adult patients: current perspectives. Adolesc Health Med Ther. 2014;5:115–25

9. Ferrari A, Dileo P, Casanova M, Bertulli R, Meazza C, Gandola L, Navarria P, Collini P, Gronchi A, Olmi P, Fossati-Bellani F, Casali PG. Rhabdomyosarcoma in adults. A retrospective analysis of 171 patients treated at a single institution. Cancer. 2003;98:571–80

10. Fischer TD, Gaitonde SG, Bandera BC, Raval MV, Vasudevan SA, Gow KW, Beierle EA, Doski JJ, Goldin AB, Langer M, Nuchtern JG, Stern S, Foshag LJ, Goldfarb M. Pediatric-protocol of multimodal therapy is associated with improved survival in AYAs and adults with rhabdomyosarcoma. Surgery. 2018;163:324–329

11. Ferrari A, Stark D, Peccatori FA, Fern L, Laurence V, Gaspar N, Bozovic-Spasojevic I, Smith O, De Munter J, Derwich K, Hjorth L, van der Graaf WTA, Soanes L, Jezdic S, Blondeel A, Bielack S, Douillard JY, Mountzios G, Saloustros E. Adolescents and young adults (AYA) with cancer: a position paper from the AYA Working Group of the European Society for Medical Oncology (ESMO) and the European Society for Paediatric Oncology (SIOPE). ESMO Open. 2021;6:100096.

12. Kahn JM, Pei Q, Friedman DL, Kaplan J, Keller FG, Hodgson D, Wu Y, Appel BE, Bhatia S, Henderson TO, Schwartz CL, Kelly KM, Castellino SM. Survival by age in paediatric and adolescent patients with Hodgkin lymphoma: a retrospective pooled analysis of children’s oncology group trials. Lancet Haematol. 2022;9:e49–e57

13. Lin, D. Y., Wei, L. J., and Ying, Z. (1993). “Checking the Cox Model with Cumulative Sums of Martingale-Based Residuals.” Biometrika 80:557–572.

14. Saloustros E, Stark DP, Michailidou K, Mountzios G, Brugieres L, Peccatori FA, Jezdic S, Essiaf S, Douillard JY, Bielack S. The care of adolescents and young adults with cancer: results of the ESMO/SIOPE survey. ESMO Open. 2017;2:e000252

15. Keegan TH, Ries LA, Barr RD, Geiger AM, Dahlke DV, Pollock BH, Bleyer WA; National Cancer Institute Next Steps for Adolescent and Young Adult Oncology Epidemiology Working Group. Comparison of cancer survival trends in the United States of adolescents and young adults with those in children and older adults. Cancer. 2016;122:1009–16

16. Spreafico F, Ferrari A, Mascarin M, Collini P, Morosi C, Biasoni D, Biassoni V, Schiavello E, Gandola L, Gattuso G, Chiaravalli S, Massimino M. Wilms tumor, medulloblastoma, and rhabdomyosarcoma in adult patients: lessons learned from the pediatric experience. Cancer Metastasis Rev. 2019;38:683–694

17. Parham DM, Barr FG. Classification of rhabdomyosarcoma and its molecular basis. Adv Anat Pathol. 2013;20:387–97

18. Malempati S, Weigel BJ, Chi YY, Tian J, Anderson JR, Parham DM, Teot LA, Rodeberg DA, Yock TI, Shulkin BL, Spunt SL, Meyer WH, Hawkins DS. The addition of cixutumumab or temozolomide to intensive multiagent chemotherapy is feasible but does not improve outcome for patients with metastatic rhabdomyosarcoma: A report from the Children’s Oncology Group. Cancer. 2019;125:290–297

19. Abraham IE, Rauscher GH, Patel AA, Pearse WB, Rajakumar P, Burkart M, Aleem A, Dave A, Bharadwaj S, Paydary K, Acevedo-Mendez M, Goparaju K, Gomez R, Carlson K, Tsai SB, Quigley JG, Galvin JP, Zia M, Larson ML, Berg S, Stock W, Altman JK, Khan I. Structural racism is a mediator of disparities in acute myeloid leukemia outcomes. Blood 2022;139:2212–2226

20. Munnikhuysen SR, Ekpo PA, Xue W, Gao Z, Lupo PJ, Venkatramani R, Heske CM. Impact of race and ethnicity on presentation and outcomes of patients treated on rhabdomyosarcoma clinical trials: A report from the Children’s Oncology Group. Cancer Med. 2023 Jun;12(11):12777–1279

21. Chen J, Liu X, Lan J, Li T, She C, Zhang Q, Yang W. Rhabdomyosarcoma in Adults: Case Series and Literature Review. Int J Womens Health. 2022 Mar 28;14:405–414

